# Implicit and explicit motor learning interventions have similar effects on walking speed in people after stroke: a randomized controlled single blind trial

**DOI:** 10.1101/19008797

**Authors:** Li-Juan Jie, Melanie Kleynen, Kenneth Meijer, Anna Beurskens, Susy Braun

## Abstract

**Background:** Therapists may use (more) implicit or (more) explicit motor learning approaches to facilitate motor skill learning of stroke patients. The use of implicit motor learning approaches has shown promising results in healthy populations.

**Objective:** To assess whether an implicit motor learning walking intervention is more effective compared to an explicit motor learning walking intervention delivered at home with regard to walking speed in people after stroke in the chronic phase of recovery.

**Design:** Randomized controlled single blind trial.

**Setting:** Home environment.

**Patients:** 79 people in the chronic phase after stroke (66.4 ± 11.0 years; 70.1 ± 64.3 months after stroke; walking speed 0.7 ± 0.3 m/s; Berg Balance Scale score 44.5 ± 9.5) were randomly assigned to an implicit (n=39) or explicit (n=41) group.

**Intervention:** Analogy learning was used as the implicit motor learning walking intervention, whereas the explicit motor learning walking intervention consisted of detailed verbal instructions. Both groups received nine training sessions, 30 minutes each, for a period of three weeks.

**Measurements:** The primary outcome was walking speed measured by the 10-Meter Walk Test. Outcome measures were assessed at baseline, immediate and 1-month post intervention.

**Results:** No statistically or clinically relevant differences between groups were obtained post intervention (between-group difference estimated 0.02 m/s [95% CI −0.04 to 0.08] and at follow-up (between-group difference estimated −0.02 m/s [95% CI −0.09 to 0.05], p=0,563).

**Limitations:** The treatment effects may have been diluted by “noise” accompanied with research within real life settings, complex tasks and a representative sample.

**Conclusions:** Implicit motor learning was not superior to the explicit motor learning to improve walking speed in people after stroke in the chronic phase of recovery.

## INTRODUCTION

One of the most practiced motor skills in stroke rehabilitation is walking^1^. In general, therapists use (more) implicit or (more) explicit forms of learning to facilitate improvement of gait. Explicit motor learning can be referred to as a more conscious form of learning, that is characterized by the generation of verbal knowledge (i.e. facts and rules about movement performance) and involvement of cognitive resources^2^. In contrast, implicit motor learning is assumed to take place without much knowledge of the underlying facts and rules of motor skills and has been described as ‘*learning that progresses with no or minimal increase in the verbal knowledge of movement performance and without awareness*’^2(p2)^. Within current clinical practice therapists tend to structure therapy in a more explicit manner or switch between implicit and explicit learning approaches^3–5^. However, this might not always be efficient. For people after stroke, who often experience cognitive impairments^6^, it can be difficult to process large amounts of verbal explicit information. Implicit motor learning, on the other hand, strives to minimize the involvement of cognitive resources, especially working memory^7^ and may therefore be more feasible for people after stroke who apart from physical constraints also suffer from cognitive impairments. Studies show that people after stroke are able to learn implicitly and that performance of an implicitly learned task might be more stable under dual-task condition and more durable over time^8^. However, there is still a lack of studies comparing the effects of implicit motor learning post-stroke to explicit motor learning within clinically relevant tasks. In order to be clinically meaningful, implicit and explicit motor learning approaches need to be tailored to the individual needs of the patients and performed in the real-life situations.

One practical approach to induce implicit motor learning is through the use of analogies. In analogy learning, the learner is provided with one single metaphor (or analogy) that strives to encompass all underlying (explicit) knowledge that is necessary to complete the motor skill. For example, to facilitate step length a therapist could provide the analogy ‘*Walk as if you follow the footprints in the sand’*^9^. Although no technical (explicit) instructions are given, the analogy may facilitate for example a more symmetrical gait, the foot strike from heel to toe and foot-clearance. Studies in athletes have shown that analogy learning led to better and more stable performance under dual-task conditions^10,11^. Within the neurological population first pilot studies reveal the feasibility of analogy learning and demonstrate its potential as both clinically relevant and statistically significant changes in walking performance could be obtained^9,12,13^. In the current study, the effects of analogy learning were compared to detailed verbal instructions when training the clinically relevant task ‘walking’ in a real life setting (home environment).

To our knowledge, this is the first randomized controlled trial that examines the effects of implicit motor learning facilitated by analogies compared to explicit motor learning on a functional walking task in people after stroke. Contrary to earlier studies examining implicit motor learning using the same analogy for the entire group^11^ the current study also tailored the interventions towards the individual needs, preferences and abilities of the patients. The research question was: Is a 3-week implicit motor learning walking intervention (analogies) more effective compared to a 3-week explicit motor learning walking intervention (verbal detailed instructions) delivered at home with regard to walking speed in people after stroke who are in the chronic phase of recovery? Walking speed was chosen due its integrated results on other gait parameters e.g. step length^14^ and functional outcomes^15^. It was hypothesized that implicit motor learning would result in greater improvements of walking speed post intervention (especially at 1-month post intervention).

## METHOD

### Study design and participants

The study adopts a randomized, controlled, single-blinded study design and was approved by the local ethics committee METC-Z in Heerlen, the Netherlands (approval number 17-T-06, Netherlands Trial Register: NL6133). Full details of the study protocol have been published elsewhere^16^. Recruitment of participants took place via community practices, rehabilitation institutes in the region and through a local health-related newspaper. Participants were included if they were > 6 month after stroke, had a self-selected walking speed lower than 1.0 m/s, were able to communicate in Dutch and to complete a three-stage command. Participants were excluded if they were unable to walk a minimum distance of 10 meter, could not ambulate on level surfaces without manual contact of another person (Functional Ambulation Scale (FAC) < 3), had additional impairments not related to stroke that significantly influenced their gait pattern (e.g. Parkinson’s disease). All participants signed a written informed consent.

### Randomization and masking

A randomization list was generated using a web-based randomization program and was only available to an independent researcher, not involved in the delivery of the interventions or measurements. Patients were randomly assigned (1:1) to either the implicit or explicit motor learning condition (block size of four). The assessors were blind to the treatment allocation. The therapists were aware of the treatment condition they provided. Patients were not told which condition they received and were asked to not reveal details about the treatment to the blinded assessors.

### Interventions

In total nine training sessions were provided over a three-week long intervention period. Each session lasted 30 minutes. Within a case-study this duration and frequency of sessions were sufficient to result in clinically meaningful changes^13^. An intervention guideline outlining how the implicit and explicit motor learning intervention should be delivered was developed for therapists in the trial. The guideline was developed with physiotherapists and client representatives and was based on the previous pilot studies and experiences^9,13,16^. Prior to the trial, five standardization training sessions with the therapists took place to discus and explicate the intervention guideline with example cases. In both interventions, therapist examined the participants walking pattern and defined the underlying gait parameters which could potentially influence walking speed. More details about the interventions and main characteristics with regard to instructions and feedback are described in figure 1^16^.

**Figure 1.**
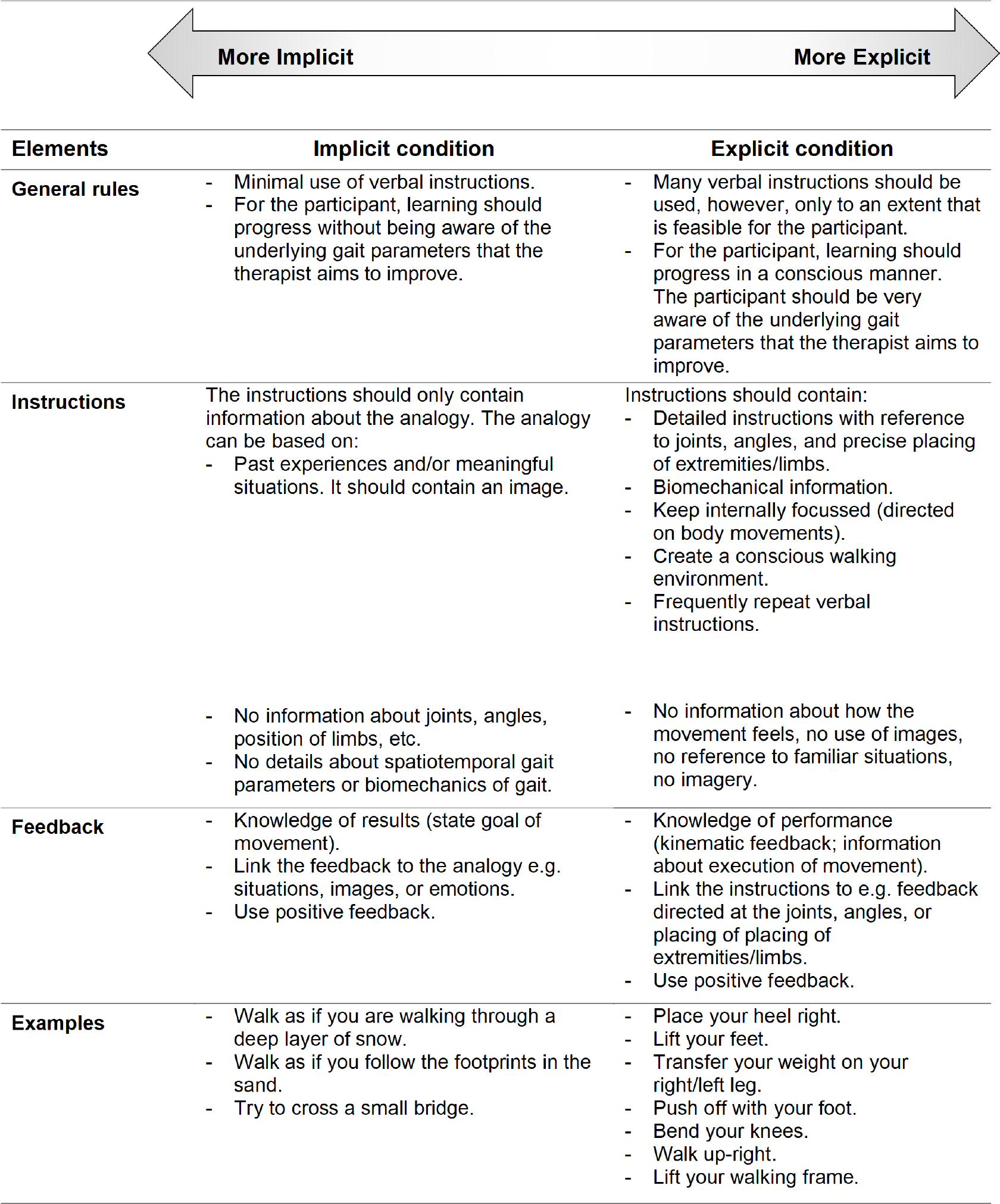
Adapted from Jie et al^16^ under the terms of Creative Commons Attribution 4.0 license

#### The implicit intervention

The main focus for the implicit intervention was creating a learning situation in which the learner was not (or minimally) aware of the underlying rules of the practiced motor skill. The concept of analogy learning formed the basis to guide the implicit intervention because 1) it has shown to adopt characteristics of implicit learning^10^ and 2) it offers therapists a practical and feasible tool to apply therapy^12,13^. The participants were provided with an analogy which aimed to improve the walking performance and was meaningful to them.

#### The explicit intervention

The main focus for the explicit intervention was creating a learning situation in which the learner is very aware of the learning process, e.g. in which he/she can precisely explicate the underlying facts and rules that are necessary to perform the motor skill. Therefore, the participant was provided with detailed explicit instructions on their gait performance.

### Outcomes

#### Demographic information

The following demographic information and clinical characteristics were collected: age, gender, time post stroke, affected side, use of walking aids, educational level, cognitive level (Montreal Cognitive Assessment, MoCA)^17^, static balance and fall risk (Berg Balance Scale)^18^, mobility disability (Rivermead Mobility Index)^19^, and ability to make movements outside the synergetic patterns (Fugl-Meyer assessment of the lower limb)^20^. To assess the propensity for conscious motor processing, the Dutch version of Movement Specific Reinvestment Scale (MSRS)^21,22^ was used.

#### Outcome assessment

The primary outcome measure was walking speed which was calculated by measuring the walking time on a 10-Meter Walk Test (10MWT; 10m / time (s))^23^. Secondary outcomes measures were the modified Dynamic Gait Index (DGI)^24^, motor and cognitive Dual Task (DT) performance^25^, Movement Specific Reinvestment Scale (MSRS) adapted for gait^16,26^, verbal protocol, Stroke and Aphasia Quality of Life Scale (SAQOL-39)^27^, Global Perceived Effect (GPE) scale^28^ and verbal protocol.

#### Assessment of the dual task

Motor task performance was measured through calculating the Dual Task Effects (DTE) according to the formula of Kelly et al^29^. The cognitive task performance was measured via calculating the error scores as done before by Wilson et al^25^. The error scores were not yet relative to single task. Therefore, the dual task error scores were subtracted from the single task error scores. Both the motor and cognitive task performances were expressed in percentages. Negative percentages indicate that performance deteriorated relative to single task, whereas positive scores indicate relative improvements of the dual task performance.

#### Verbal Protocol

To assess the amount of explicit knowledge, a verbal protocol questionnaire was be administered after the three-week intervention^8^. Explicit knowledge is assessed by examining the number of explicit rules that the participant used during walking. More information of the definition of ‘explicit rule’ is described elsewhere^16^. The answers of the verbal protocol were screened by two independent researchers who were blind to the experimental intervention.

### Sample size calculation and statistical analyses

The sample size calculation resulted in a minimum group size of 33 participants per group. The power was set at beta 0.80, the significance level at alpha 0.05, and a standard error of 0.23m/s^30^. The minimal clinically important difference (MCID) of 0.16 m/s for walking speed was set as the minimal change^31^. Considering 10% of participants may be lost during (drop-out) and another 10% after the intervention (loss-to-follow-up), this study aimed to recruit 40 participants per group. The statistical analyses were performed using IBM SPSS (version 24). Baseline characteristics of the two groups were reported using frequency distributions and descriptive statistics. For the *intention-to-treat analyses* data of all participants who received the intervention were analysed according to their original treatment allocation. Treatment effects on numerical data were assessed using a linear mixed model. The model represented group, time and group × time as fixed factors. For the repeated measures (balanced design) an unstructured covariance structure was used. The linear mixed model analyses uses all available data, corrects for baseline differences and accounts for dependency of data.

Statistical analyses of the primary outcome was also described in relation to clinically relevant differences between groups (MCID: 0.16 m/s)^31^. In the *per-protocol analyses* data of subjects were excluded if they did not receive the intervention as intended i.e. when protocol deviations occurred in two or more (of the nine) sessions. Possible protocol deviations were self-reported (subjective) in therapists logs and randomly 10 gait training sessions were audio-recorded (objective) and evaluated to detect protocol deviations. Furthermore, people who did not meet the inclusion criteria or people who dropped out were excluded in the per-protocol analysis. Descriptive sub-group analysis was performed on cognition to explore whether cognitive abilities (MoCA ≤ 21) might influence the effect of the interventions. The verbal protocol was only assessed once and an independent t-test was used to compare results between the groups.

## RESULTS

### Flow of participants through the trial

The flowchart of the trial is presented in figure 2. Between 19 May 2017 and 19 September 2018, a total 81 people were assessed for eligibility and randomized. Two participants (3%) did not start with the study. One participant withdrew due to diagnoses with additional impairments that severely influenced his gait. The other participant decided to stop due to personal reasons. All participants (n=79) that started the intervention were included in the primary intention-to-treat analysis. Demographics and baseline characteristics are presented in table 1. There were no apparent differences between the groups at baseline.

**Table 1.** Baseline characteristics of participants

|  | Implicit<br>(n = 38) | Explicit<br>(n = 41) |
| --- | --- | --- |
| **General Characteristics** |  |  |
| Age (yr), mean (SD) | 64.6 (9.4) | 67.8 (11.6) |
| Gender, n males (%) | 24 (63%) | 25 (61%) |
| Length (cm), mean (SD) | 171.7 (8.0) | 172.0 (8.8) |
| Educational level, n (%) |  |  |
| Elementary education | 0 (0%) | 1 (2%) |
| Secondary education | 20 (53%) | 18 (44%) |
| Vocational training | 10 (26%) | 11 (27%) |
| University | 8 (21%) | 11 (27%) |
| **Stroke Characteristics** |  |  |
| Time post stroke (months), mean (SD) | 72.8 (59.3) | 67.5 (69.1) |
| Side of stroke, n right (%) | 19 (50%) | 16 (19%) |
| Independent walking, n (%) |  |  |
| Walk unaided | 9 (24%) | 9 (22%) |
| Walk with stick | 29 (67%) | 32 (78%) |
| **Motor Characteristics** |  |  |
| Berg Balance Scale (0-56), mean (SD) | 45.5 (11.6) | 43.5 (8.9) |
| Rivermead Mobility Index (0-15), mean (SD) | 11.6 (2.5) | 11.3 (2.7) |
| Fugl-Mayer Assessment (0-34), mean (SD) | 23.5 (8.0) | 22.2 (8.1)* |
| **Cognitive Characteristics** |  |  |
| Montreal Cognitive Assessment (points/30, mean (SD) | 24.7 (4.2) | 23.2 (6.2) |
| **Conscious motor control preference** |  |  |
| Movement Specific Reinvestment Scale (0-10), mean (SD) | 4.9 (2.5) | 5.1 (2.6) |

\* Due to fatigue one participant from the explicit group was unable to complete the Fugl-Mayer Assessment

**Figure 2.**
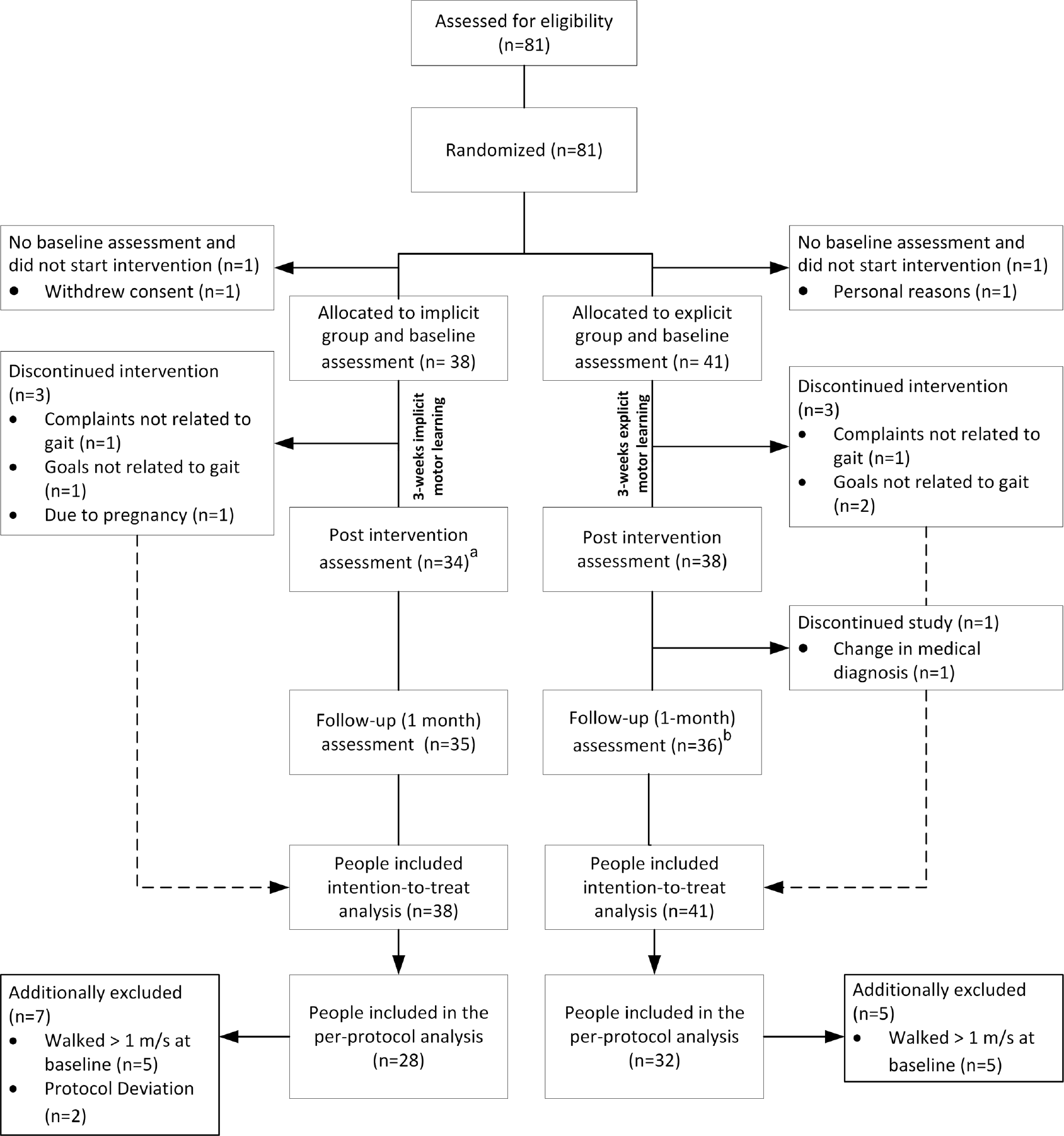
Flowchart of the trial. ^a^ One participant was unavailable for the post intervention assessment (n=1) ^b^ One participant was unavailable for the follow-up assessment (n=1)

### Compliance with the trial

Two participants (3%) deviated from the protocol with regard to the provided instructions (>2 explicit instructions within the implicit intervention). Analysis revealed that in retrospect ten participants (13%) did not meet the inclusion criteria of walking slower than 1 m/s at baseline. In addition, three participants (4%) wanted to improve overall fitness but had no specific goals related to gait and therefore discontinued with the intervention. Two participants (3%) stopped due to other complaints not related to gait. Furthermore, the medical diagnoses of one participant (initially stroke; 1%) was changed during the intervention. Due to pregnancy another participant dropped out of the intervention. All available data of these 19 participants (24%) were included in the primary intention-to-treat analysis but were excluded in the per-protocol analysis.

### Results of the intention-to-treat analysis

Table 2 presents the observed means (SD) per group and time point, the within-group differences, and the estimated between-group differences of the implicit versus the explicit group. Mixed linear models revealed no statistically significant differences between the groups immediately after (difference estimate 0.02 m/s [95% CI −0.04 to 0.08], p=0.498) and one-month post intervention (difference estimate −0.02 m/s [95% CI −0.09 to 0.05], p=0,563; see table 2 and figure 3). Also, no clinically relevant (MCID: 0.16 m/s^31^) difference between groups were observed. No statistically significant differences in favour of any group was obtained on any of the other secondary outcome parameters (table 2). Over time, within groups, positive changes were observed in outcome measures related to gait function (DGI, DTmotor), and quality of life (SAQOL-39). Participants’ perceived effects with regard to the intervention was similar for both groups (see figure 4). With regard to the verbal protocol, on average, people in the implicit group accumulated significantly fewer explicit rules (*M*=0.38, *R*= 0 to 2 rules, *SE*=0.10) compared to the explicit group (*M*=2.42, *R*= 0 to 6 rules, *SE*=0.27), t(68)=-7.07, p<.05 after the intervention.

**Table 2.**
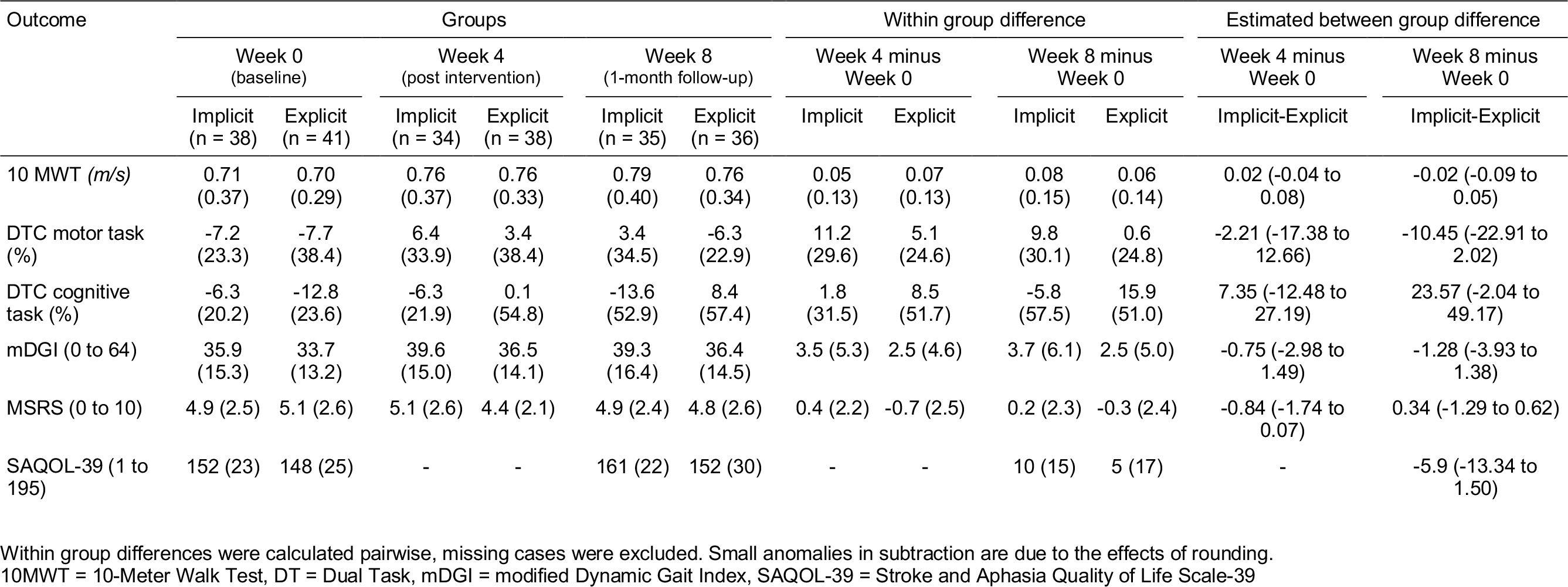
Mean (SD) of groups, mean (SD) difference within groups, and estimated mean (95% CI) difference between groups as established with linear mixed model.

| Outcome | Groups |  |  |  |  |  | Within group difference |  |  |  | Estimated between group difference |  |
| --- | --- | --- | --- | --- | --- | --- | --- | --- | --- | --- | --- | --- |
|  | Week 0<br>(baseline) |  | Week 4<br>(post intervention) |  | Week 8<br>(1-month follow-up) |  | Week 4 minus<br>Week 0 |  | Week 8 minus<br>Week 0 |  | Week 4 minus<br>Week 0 | Week 8 minus<br>Week 0 |
|  | Implicit<br>(n = 38) | Explicit<br>(n = 41) | Implicit<br>(n = 34) | Explicit<br>(n = 38) | Implicit<br>(n = 35) | Explicit<br>(n = 36) | Implicit | Explicit | Implicit | Explicit | Implicit-Explicit | Implicit-Explicit |
| 10 MWT (*m/s*) | 0.71<br>(0.37) | 0.70<br>(0.29) | 0.76<br>(0.37) | 0.76<br>(0.33) | 0.79<br>(0.40) | 0.76<br>(0.34) | 0.05<br>(0.13) | 0.07<br>(0.13) | 0.08<br>(0.15) | 0.06<br>(0.14) | 0.02 (-0.04 to<br>0.08) | -0.02 (-0.09 to<br>0.05) |
| DTC motor task<br>(%) | -7.2<br>(23.3) | -7.7<br>(38.4) | 6.4<br>(33.9) | 3.4<br>(38.4) | 3.4<br>(34.5) | -6.3<br>(22.9) | 11.2<br>(29.6) | 5.1<br>(24.6) | 9.8<br>(30.1) | 0.6<br>(24.8) | -2.21 (-17.38 to<br>12.66) | -10.45 (-22.91 to<br>2.02) |
| DTC cognitive<br>task (%) | -6.3<br>(20.2) | -12.8<br>(23.6) | -6.3<br>(21.9) | 0.1<br>(54.8) | -13.6<br>(52.9) | 8.4<br>(57.4) | 1.8<br>(31.5) | 8.5<br>(51.7) | -5.8<br>(57.5) | 15.9<br>(51.0) | 7.35 (-12.48 to<br>27.19) | 23.57 (-2.04 to<br>49.17) |
| mDGI (0 to 64) | 35.9<br>(15.3) | 33.7<br>(13.2) | 39.6<br>(15.0) | 36.5<br>(14.1) | 39.3<br>(16.4) | 36.4<br>(14.5) | 3.5 (5.3) | 2.5 (4.6) | 3.7 (6.1) | 2.5 (5.0) | -0.75 (-2.98 to<br>1.49) | -1.28 (-3.93 to<br>1.38) |
| MSRS (0 to 10) | 4.9 (2.5) | 5.1 (2.6) | 5.1 (2.6) | 4.4 (2.1) | 4.9 (2.4) | 4.8 (2.6) | 0.4 (2.2) | -0.7 (2.5) | 0.2 (2.3) | -0.3 (2.4) | -0.84 (-1.74 to<br>0.07) | 0.34 (-1.29 to 0.62) |
| SAQOL-39 (1 to<br>195) | 152 (23) | 148 (25) | - | - | 161 (22) | 152 (30) | - | - | 10 (15) | 5 (17) | - | -5.9 (-13.34 to<br>1.50) |

Within group differences were calculated pairwise, missing cases were excluded. Small anomalies in subtraction are due to the effects of rounding. 10MWT = 10-Meter Walk Test, DT = Dual Task, mDGI = modified Dynamic Gait Index, SAQOL-39 = Stroke and Aphasia Quality of Life Scale-39

**Figure 3.**
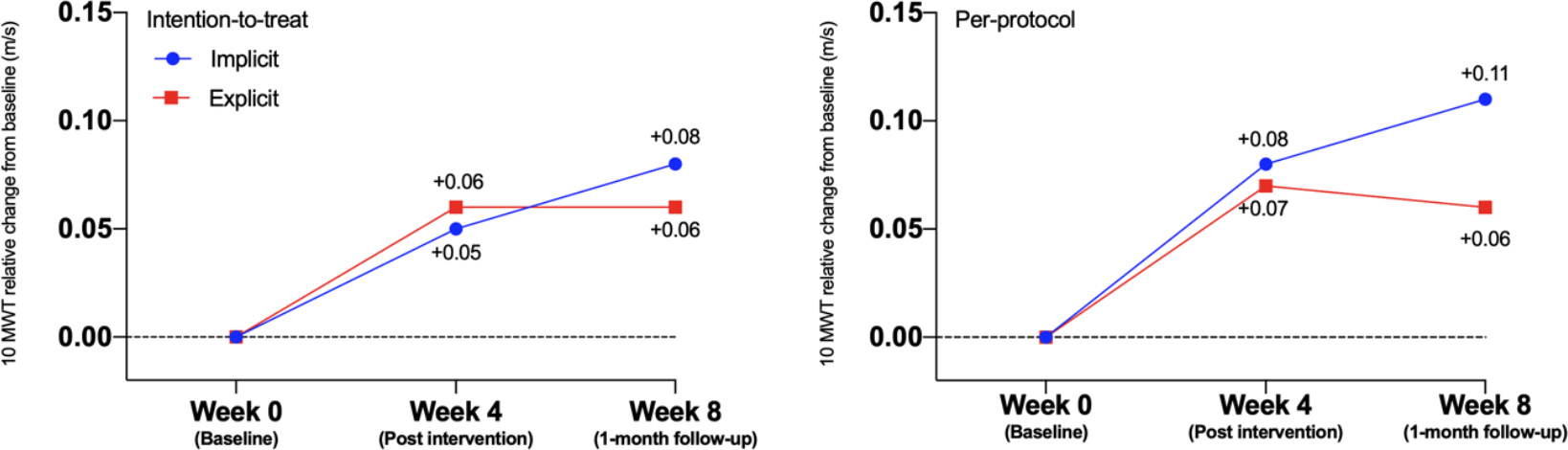
Performance of the 10 Meter Walking Test

**Figure 4.**
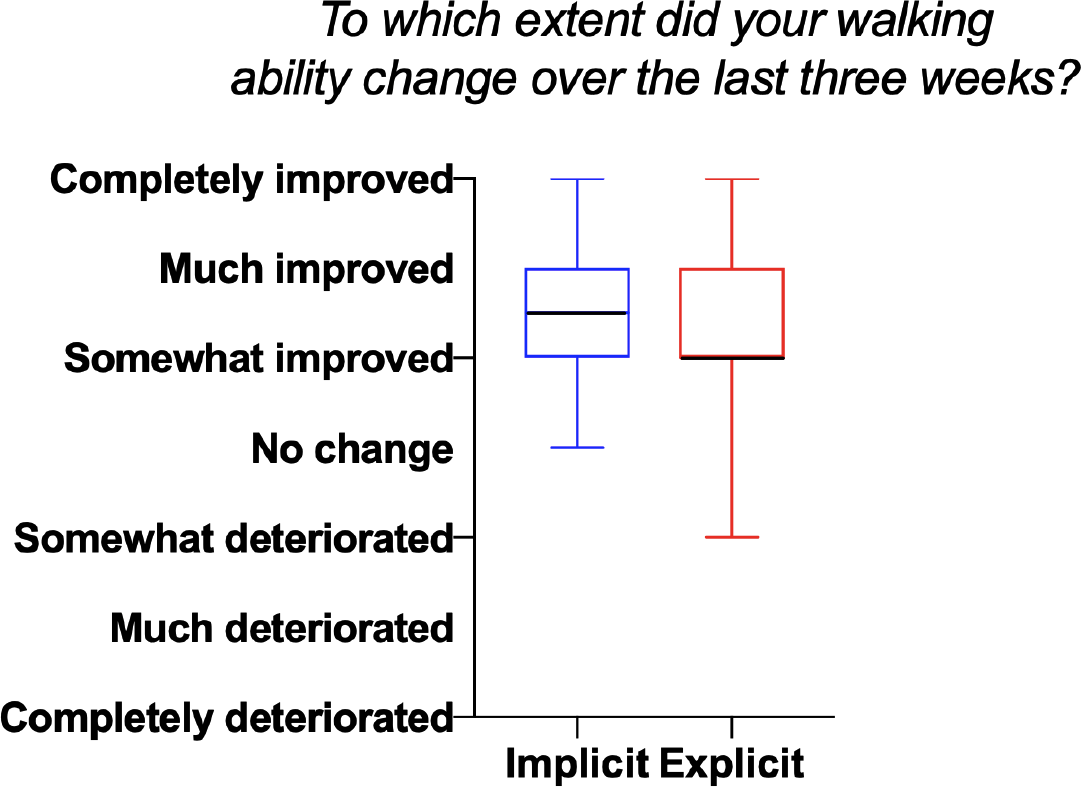
Boxplot of the Global Perceived Effect

#### Sub-group analysis on cognition

In total 15 people (implicit group n=5; explicit group n=10) had a MoCA score < 22. No trend in favour of the implicit intervention was observed in the descriptive sub-group analysis on cognition (see supplementary data 1).

### Results of the Per-Protocol

The per-protocol analyses led to slightly larger changes between groups but again did not lead to statistically significant after (difference estimate −0.06 m/s [95% CI −0.13 to 0.02], p=0.140) or clinically relevant effects (MCID: 0.16 m/s) between group on the primary outcome (see lower graph figure 3 and supplementary data 2).

## DISCUSSION

People after stroke in the chronic phase of recovery who received an implicit motor learning walking intervention (analogies) under the guidance of physiotherapists in their home environments, had similar effects on walking speed compared to those who received an explicit motor learning walking intervention (verbal detailed instructions). No statistically nor clinically relevant differences between groups were found as between group differences (intention-to-treat Δ 0.02 m/s; per-protocol Δ 0.05 m/s) did not exceed the chosen clinically relevant threshold of 0.16 m/s^31^. Similarly, based on the descriptive sub-group analysis on cognition (MoCA <22) no trend in favour of the implicit intervention was observed (table 3). No statistical sub-group analysis on cognition was performed based on small group sizes.

To our knowledge this was the first and largest trial in the field of stroke rehabilitation to examine the effectiveness of implicit motor learning to improve the functional ‘walking’ task within a clinically relevant context (home environment of the patient)^32^. The results of this study did not replicate the more promising findings on implicit motor learning in stroke from earlier studies, generally performed in more standardized, laboratory settings and/or with non-functional tasks e.g.^33–35^. A variety of factors related to the selection of participants (selection bias), use of the 10MWT as primary outcome measure (information bias) and operationalization of the intervention (contrasts) may have influenced the results and led to these neutral findings.

First, a selection bias may have occurred. To increase generalizability of the results and to gain a better insight into the potential effects in clinical practice, we chose to include a sample of stroke patients, which reflects the heterogeneity of the stroke population as seen in rehabilitation. The researched target population group therefore showed a large variability in terms of demographics as well as physical and cognitive abilities. This heterogeneity may reflect reality in practice but might also have diminished the results. Further, the erroneous inclusion of ten participants whose baseline walking speed exceeded the inclusion criterion may have led to a ceiling effect. This ceiling effect might explain the larger, but not significant, trend towards implicit motor learning (see figure 3 lower graph) in the per-protocol analysis. In addition, the study was probably underpowered due to this deviation.

Second, the use of the 10MWT as primary outcome measure may have implications for both the findings themselves and the interpretation in terms of clinical meaningfulness. The 10MWT was chosen as the primary outcome measure due to its validity, reliability and feasibility within clinical practice^36^ but also to allow comparison with other studies^37^. The advantage of using walking speed as a primary outcome is the integrated result on multiple gait parameters such as step length and frequency^14^,^15^ and the direct relation to changes in functional scales^38^. However, it could be argued that the 10MWT might not have been sensitive enough to detect changes due to implicit learning if those underlying changes are small or not obviously related to walking speed (e.g. improvement of confidence during walking).

Another explanation for the neutral results could originate from the way the interventions were operationalised. Contrary to earlier studies in more controlled settings and with non-functional tasks such as serial reaction time tasks^32^, it seems difficult to keep the contrast between interventions equally large when including a functional task within a clinically relevant environment. In contrast to other studies^10,11^ the exact number of rules were not pre-defined but tailored to the participants. For example, Lam et al used a fixed number of ‘eight’ verbal rules compared to ‘one’ analogy^11^. The provided number of explicit rules (explicit intervention) may have been limited because of ethical reasons, potentially resulting in a diminished contrast between groups.

Within this study we assessed the implicit nature of the intervention by asking participants to report the number of explicit rules the learned (verbal protocol), assessing durability of performance over a longer time period and dual task interference^10,11^. None of these measures revealed a clear picture on the nature of the learning process. For instance, fewer rules were accumulated in the implicit compared to the explicit group, but it remains unclear whether these rules have been acquired through treatments before enrolment of this study. In addition, for some participants the tone-counting task may have been too easy not leading to dual task interference, whereas for other people the task was too difficult. Due to this large variation in performance on the cognitive (tone counting) dual task it was not possible to further legitimately interpret these results.

Finally, on average both groups slightly improved their walking speed after the intervention (+0.08 m/s in the implicit group and +0.06 m/s in the explicit group) exceeding the threshold for clinical relevant change of > 0.06 m/s for within group differences as established by Perera et al^39^. It might be that using implicit or explicit motor learning does not make a (clinically relevant) difference for the results of walking rehabilitation within the included target group and setting of this trial. It is remarkable that the detected improvement (in both groups) remained relatively stable at the follow-up test. This finding might be seen as a form of retention and indicates that motor learning occurred rather than just a temporal improvement in motor performance.

## Future research

The design of this RCT was carefully prepared by research into underlying theories^40^, feasibility and piloting testing^12,13^ of implicit motor learning^41^. Applying the RCT in its cleanest form in clinical settings^42^ and with complex interventions was challenging as we needed to balance between external validity (generalizability of the results for daily practice) and internal validity (standardization and reliability of the results). Other designs may be considered to evaluate effectiveness of long-term, highly individualized, and complex interventions^41^, as needed in the field of motor learning. Two recent studies suggest that tailoring motor learning interventions towards patient characteristics and preferences might be important, promoting more pragmatic trials^9,43^. The interventions may also be applicable for people with more severe cognitive impairments (MoCA <21) as equal trends in performance were found within this sub-group. A logical next step would be to assess which patient characteristics influence motor learning interventions and how these factors influence the learning process. Therefore, cohort studies in which all potential influencing factors (e.g. activity dependent plasticity, cognition, or individual preferences) are measured over time and therapist document the used motor learning approach in detail might be an interesting alternative to consider.

To gain more insight in the gait mechanisms and functional effects when applying implicit motor learning, future studies may consider combining upcoming instruments for quantitative gait analysis which can be performed outside laboratory settings (e.g. use of wearable sensors^44,45^) with patient specific outcome measures which can detect functional relevant changes within individualized goals (e.g., Patient Specific Functional Scale)^46,47^.

## Conclusion and clinical message

In this study, no overall benefits of implicit motor learning over explicit motor learning for improving walking performance in people after stroke in the chronic phase of recovery were found. The treatment effects in this study may have been diluted by “noise” accompanied with research within real life settings, complex tasks and a representative sample of the target population. For tailored motor learning approaches more insight is needed on the patient characteristics and preferences that influence the process of motor learning. While awaiting further results, therapists may consider both motor learning approaches to facilitate walking speed within the stroke population.

## Data Availability

The data that support the findings of this study are available from the corresponding author, upon reasonable request.

## Acknowledgements

The authors would like to thank the therapists, and researchers who contributed in conducting the trial: Jos Halfens, Jos Kurvers, Wiel Regtop, Kyra Theunissen, Monique Rothbauer, Inge de Lang, Ritch te Kampen, Renée van den Heuvel, Joep Westbroek. Furthermore, we thank the client participants Nathalie Sieben, Else de Bont, and Anja Minheere for their perspectives and thoughts on the trial during the set-up of the study. We thank Peter Konsten for his support on the assessment forms and Bjorn Winkens for his advice in the data analyses. Finally, we thank all participants that took part in the study.

